# The landscape of COVID-19 vaccination among healthcare workers at the first round of COVID-19 vaccination in China: willingness, acceptance and self-reported adverse effects

**DOI:** 10.1101/2021.05.15.21257094

**Authors:** Xinxin Ye, Wan Ye, Jinyue Yu, Yuzhen Gao, Ziyang Ren, Lanzhen Chen, Ao Dong, Qian Yi, Chenju Zhan, Yanni Lin, Yangxin Wang, Simin Huang, Peige Song

## Abstract

**Background:** The COVID-19 vaccines have been developed in a wide range of countries. This study aims to examine factors influencing vaccination rate and willingness to vaccinate against COVID-19 among Chinese healthcare workers (HCWs).

**Methods:** From 3rd February to 18th February, 2021, an online cross-sectional survey was conducted among HCWs to investigate factors associated with the acceptance and willingness of COVID-19 vaccination. Socio-demographic characteristics and the acceptance of COVID-19 vaccination among Chinese HCWs were evaluated.

**Results:** A total of 2156 HCWs from 21 provinces in China responded to this survey (response rate: 98.99%)), among whom 1433 (66.5%) were vaccinated at least one dose. Higher vaccination rates were associated with older age (40-50 years vs. less than 30 years, OR=1.63, 95%CI: 1.02-2.58; >50 years vs. 30 years, OR=1.90, 95%CI: 1.02-3.52), working as a clinician (OR=1.54, 95% CI: 1.05-2.27), having no personal religion (OR=1.35, 95%CI: 1.06-1.71), working in a fever clinic (OR=4.50, 95%CI:1.54-13.17) or higher hospital level (Municipal vs. County, OR=2.01, 95%CI: 1.28-3.16; Provincial vs. County, OR=2.01, 95%CI: 1.25-3.22) and having knowledge training of vaccine (OR=1.67, 95%CI:1.27-2.22), family history for influenza vaccination (OR=1.89, 95%CI:1.49-2.35) and strong familiarity with the vaccine (OR=1.43, 95%CI:1.05-1.95) (All P<0.05). Strong willingness for vaccination was related to having a working in midwestern China (OR=1.89, 95%CI:1.24-2.89), considerable knowledge of the vaccine (familiar vs. not familiar, OR=1.67, 95%CI: 1.17-2.39; strongly familiar vs. not familiar, OR=2.47, 95%CI: 1.36-4.49), knowledge training of vaccine (OR=1.61, 95%CI: 1.05-2.48) and strong confidence in the vaccine (OR=3.84, 95%CI: 2.09-7.07).

**Conclusion:** Personal characteristics, working environments, familiarity and confidence in the vaccine were related to vaccination rates and willingness to get vaccinated among healthcare workers. Results of this study could provide evidence for the government to improve vaccine coverage by addressing vaccine hesitancy in the COVID-19 pandemic and future public health emergencies.

## Introduction

Severe acute respiratory syndrome coronavirus 2 (SARS-CoV-2), the virus that causes COVID-19, emerged in late 2019 and has caused a global pandemic. The pandemic has led to more than 90 million cases and 1.9 million deaths worldwide, with disastrous consequences for the world economy and public health.^1^ To achieve herd immunity and finally end the pandemic, 60 to 70% of the world population were suggested to be immune, either though natural infection or vaccination.^2^

Vaccination is one of the most effective health interventions to prevent and control the spread of infectious diseases.^1,3^ Safe and effective vaccines against SARS-CoV-2 are necessary to protect populations from COVID-19 and to safeguard global economies from continued disruption.^3,4^ The first human clinical trial of a SARS-CoV-2 vaccine (mRNA-1273) commenced on March 2020 in the United States,^5^ and a 94.1% efficacy of this vaccine has been confirmed^6^. However, the global uptake of the SARS-CoV-2 vaccine remains insufficient for herd immunity.^7,8^ To date (3^rd^ June 2021), over 681.9 million doses of SARS-CoV-2 vaccine have been administered in China, which were still under the 60% coverage as recommended. ^9^ Meanwhile, some high-risk, low-income countries, such as Afghanistan, Ethiopia, and Guinea, have not even released vaccination data.^10^ One of the reasons for the vaccine hesitancy may be the doubt about its effectiveness and safety; a survey in the United States showed that 31% of adults were not willing to get the vaccine due to a fear of side effects,^11^ and another study in France reported the 26% of adults felt resistance toward receiving the vaccine due to doubts of its effectiveness.^12^ Furthermore, a survey in China indicates the gap between people’s willingness to accept the vaccine and their actual vaccinating activity; about 47.8% of participants expressed “willingness” to receive the vaccine, but they will postpone vaccination until the safety of the vaccine is confirmed.^13^

Healthcare workers (HCWs) are high-risk groups during the COVID-19 pandemic.^14,15^ The infection risk for this group is 9-11 times higher than that of the general population.^16^ Once HCWs are infected, the infection risk for patients can consequently increase. Hence, understanding the willingness of HCWs to accept the SARS-CoV-2 vaccine and exploring the determinants for vaccinating action can help formulate targeted education and vaccine-promoting policies, which is of great importance in enhancing vaccine uptake and avoiding future outbreaks. Much of the existing literature either focuses on evaluating the explicit reasons for vaccine hesitance and resistance,^5,17,18^ or investigates the relationship between vaccination intention and sociodemographic factors of the general public by using health belief theory or planning behavior theory.^19-22^ There is a number of investigations identifying the psychological processes of people’s decision to be vaccinated and distinguishing them from those who have the intention but will not take action.

The multiple health locus of control (MHLC) scale was developed to investigate one’s beliefs that the source of reinforcements for their health-related behaviors is primarily internal (determined by their own opinion) or external (determined by a matter of chance, or under the control of powerful persons).^23^ Nowadays, the scale has been used as one of the most efficient measures for health-related behaviors.^24-26^ The present study aims to use this measurement to investigate whether the decision for accepting the COVID-19 vaccine is controlled by internal or external factors in HCWs. This study also evaluated factors influencing actual vaccination rate and willingness among HCWs in China. Results can be used to make further recommendations for corresponding vaccination strategies and immunization plans, which are of particular importance in increasing the vaccine coverage.

## Materials and Methods

### Recruitment

The inclusion criteria for this study were: (1) age ≥ 18 years old; and (2) hospital HCWs, including any doctors and nurses who worked full time at public hospitals or local clinics. All respondents gave informed consent and voluntarily participated in the survey. The exclusion criteria included (1) interns, student nurses, and medical students in school; and (2) individuals who were employed by private hospitals.

### Questionnaire

The questionnaire contained the following three parts: demographics, vaccination-related intentions and behaviors, and the MHLC scales:

1. Demographic information (13 items): participants’ gender, age, education background, religion, income, living area, field of work, time of employment, clinical occupation, and level of the hospital.
2. COVID-19 vaccination-related features: vaccination status, willingness to vaccinate, and vaccine-related knowledge.
3. The MHLC scale: The scale consists of three parts, including the internal health locus of control (IHLC, beliefs that health outcomes are related to one’s own ability and effort, Cronbach □=0.61-0.80), powerful other’s health locus of control (PHLC, beliefs that health outcomes are related to powerful others such as physicians, Cronbach □=0.56-0.75), and chance health locus of control (CHLC, beliefs that health outcomes are related to chance and fate, Cronbach □=0.55-0.83).^23,26,27^ Each part has six items (score range: 6-36), and a higher score represents a higher locus of control.^23^

### Data collection

This data collection was conducted from 3^rd^ to 18^th^ February, 2021, using a one-time anonymous online questionnaire. A pre-survey was conducted by selecting ten health professionals from different hospitals to finalize the questionnaire. The questionnaire was distributed by invitation through the social media group. Instructions were clearly provided and each questionnaire was completed with the assistance of a trained nurse. Each hospital has one or two training officers for questionnaire distribution and data collection. Details are shown in Figure A1 in Appendix.

### Statistical analysis

Statistical analysis was performed using the R Foundation for Statistical Computing (version 4.0.3). Continuous variables were reported as mean and standard deviation (SD). Dichotomous data were presented as frequency (%) and compared by Chi-square or Fisher’s exact test in two groups. Univariate and multivariate logistic regression analyses were performed to determine independent risk factors. Multivariate analysis data were represented on a forest plot for all comparative odds ratio (OR) values with 95% confidence interval (CI). P values less than 0.05 were considered statistically significant. The bar plots of some potentially related reasons were presented to analyze differences among groups. Main packages including “forest plot,” “glm,” “ggolot2,” “maps,” “map data,” and “tableone” were applied to visualize and analyze the results and make conclusions.

This study was approved by the Medical Ethics Committee of Xiamen Medical College and passed the audit of China Clinical Trial Registration Center (Registration number: ChiCTR2100042804).

## Results

### Sociodemographic characteristics of survey respondents

Between 3^rd^ February and 18^th^ February, 2021, a total of 2178 HCWs were recruited from 21 provinces across China, including 343 doctors and 1814 nurses. After removing 22 invalid questionnaires, 2156 were finally enrolled for data analysis (effective response rate: 98.99%). A total of 1433 participants were vaccinated (66.5%). Individuals were categorized as vaccinated if they had been vaccinated at least one time at the completion of the survey (Table 1). The mean age of participants was 32.91 years (SD=8.29). The sources of vaccine-related information were: work units (84.6%), WeChat (80.1%), network news (79.7%), TV (64.0%), government announcements (62.2%), community/village epidemic prevention pamphlet/bulletin board/campaign (46.4%), SMS (42.3%), other apps (35.5%), informed by others (30.7%), blogs (30.6%), and radio (10.7%) (Figure. A2).

**Table 1.**
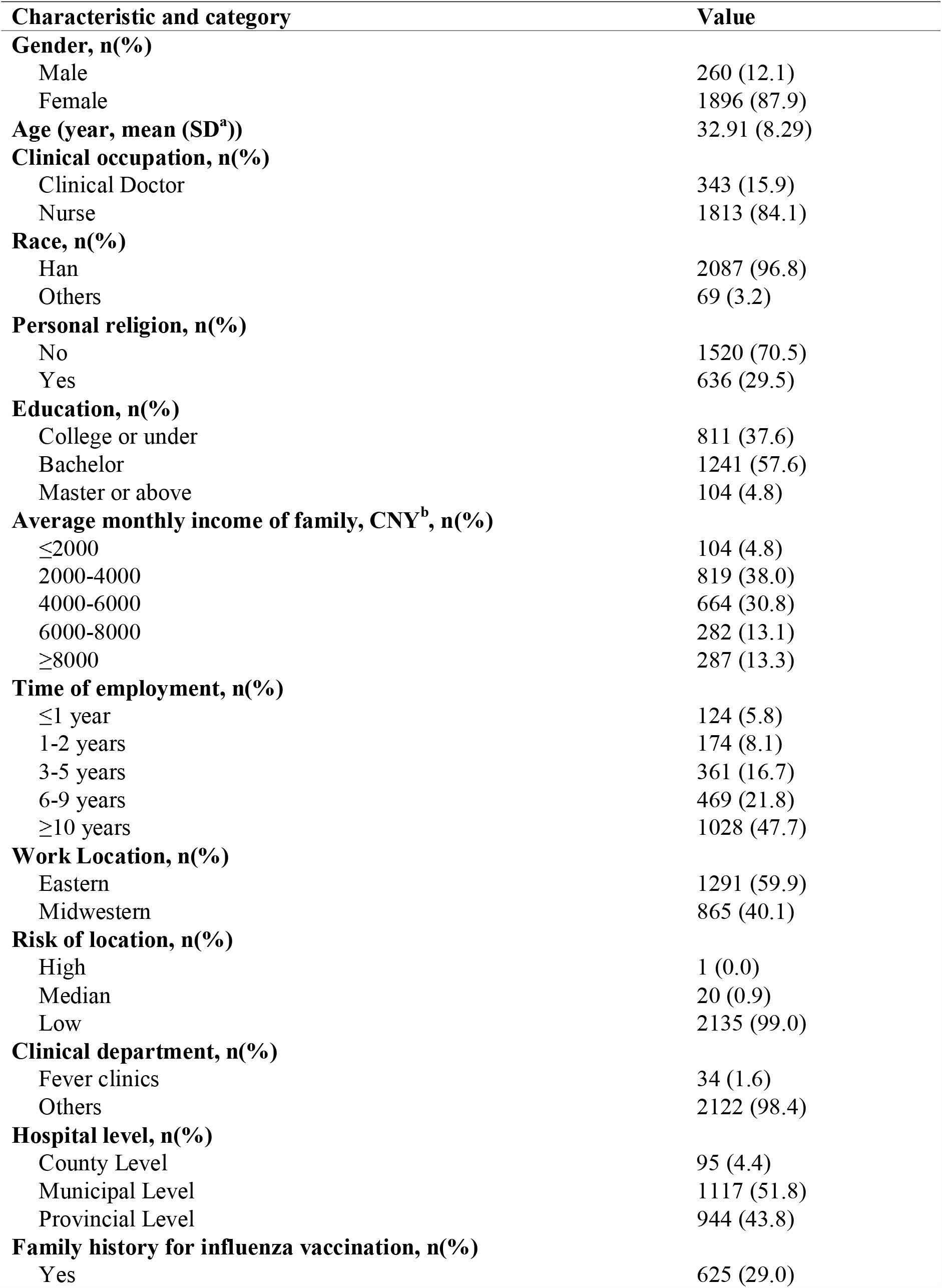
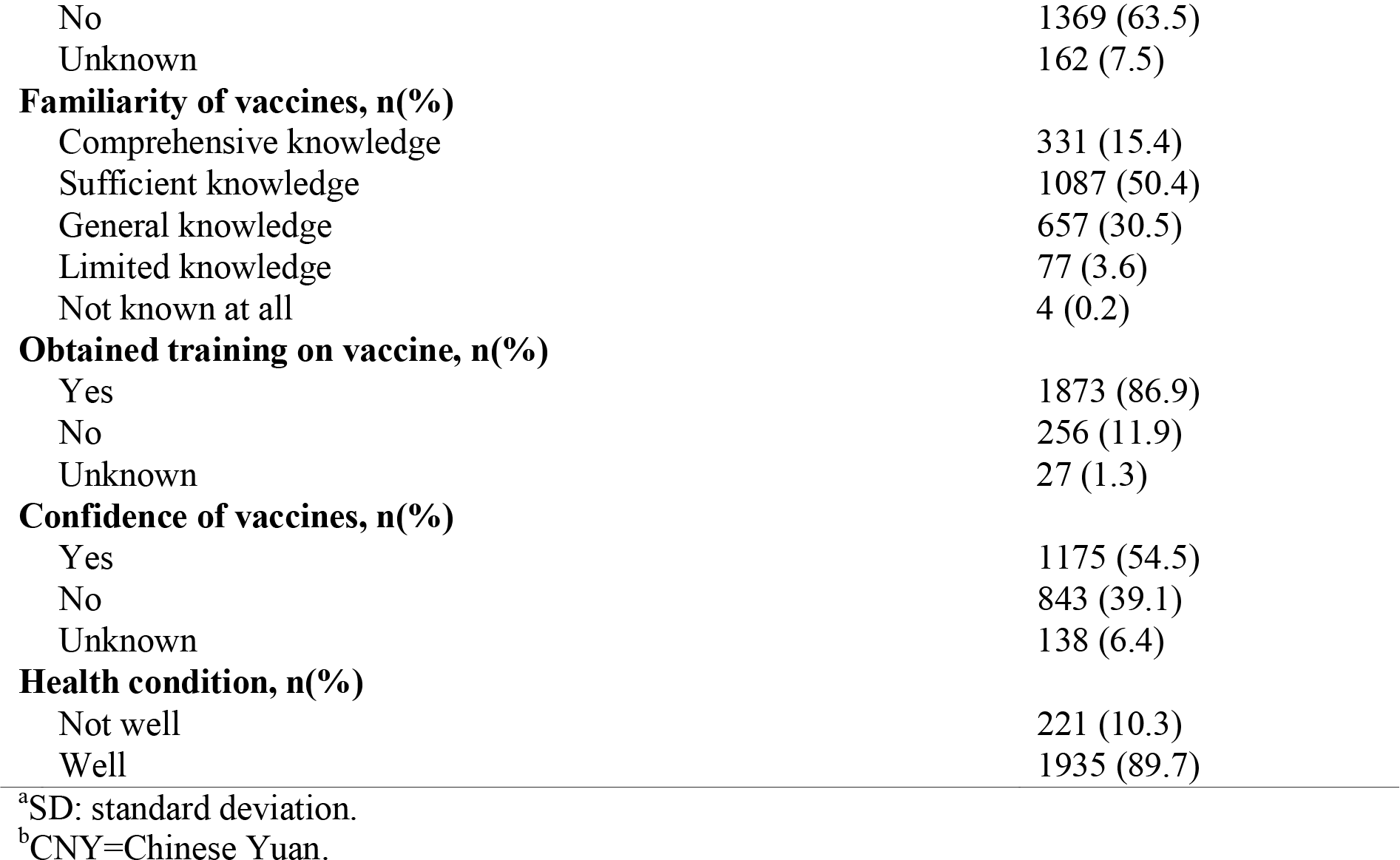
The characteristics of including subjects for vaccines in the survey

### Comparisons in people with different vaccinated status

Compared to HCWs who had not been vaccinated, vaccinated individuals were more likely to report having employment for a long time, working in midwestern China, working in a fever clinic, or a municipal hospital (Table A1). Moreover, vaccinated individuals were significantly more familiar and confident in the COVID-19 vaccines than unvaccinated HCWs. Among unvaccinated HCWs, significant differences were also observed in socio-demographics among those with different levels of willingness to vaccinate (Table A1). For instance, those who expressed a strong willingness to receive the vaccine appeared to have no personal religion, better health condition, work in midwestern China, have sufficient knowledge about the vaccine, and have strong confidence in the vaccine (all tested P<0.05).

### MHLC psychology results for the participants

To detect whether accepting the vaccine was influenced by internal or external factors, the MHLC scale was adopted. No significant difference was found in IHLC and PHLC between vaccinated and unvaccinated populations. However, the PHLC score positively related to vaccination intention in the study population (Figure A3), reflecting that subjects’ willingness to accept the vaccine may mainly be influenced by external factors, especially by powerful others, such as endorsements from the Centers for Disease Control and Prevention (CDC) in China or recommendations by physicians during the current circumstances.

### Determinants of vaccinating action and willingness of vaccination

Based on the results of the MHLC, univariate and multivariate logistic regression analyses were conducted to explore factors influencing the acceptance of vaccines in HCWs (Table A1 and Figure 1). The unvaccinated population was divided into two groups: willingness (individuals who chose “strong willingness” or “relatively strong willingness” for accepting the vaccine) and not willingness groups (individuals who chose “moderate willingness,” “prefer not,” and “not at all” for vaccination).

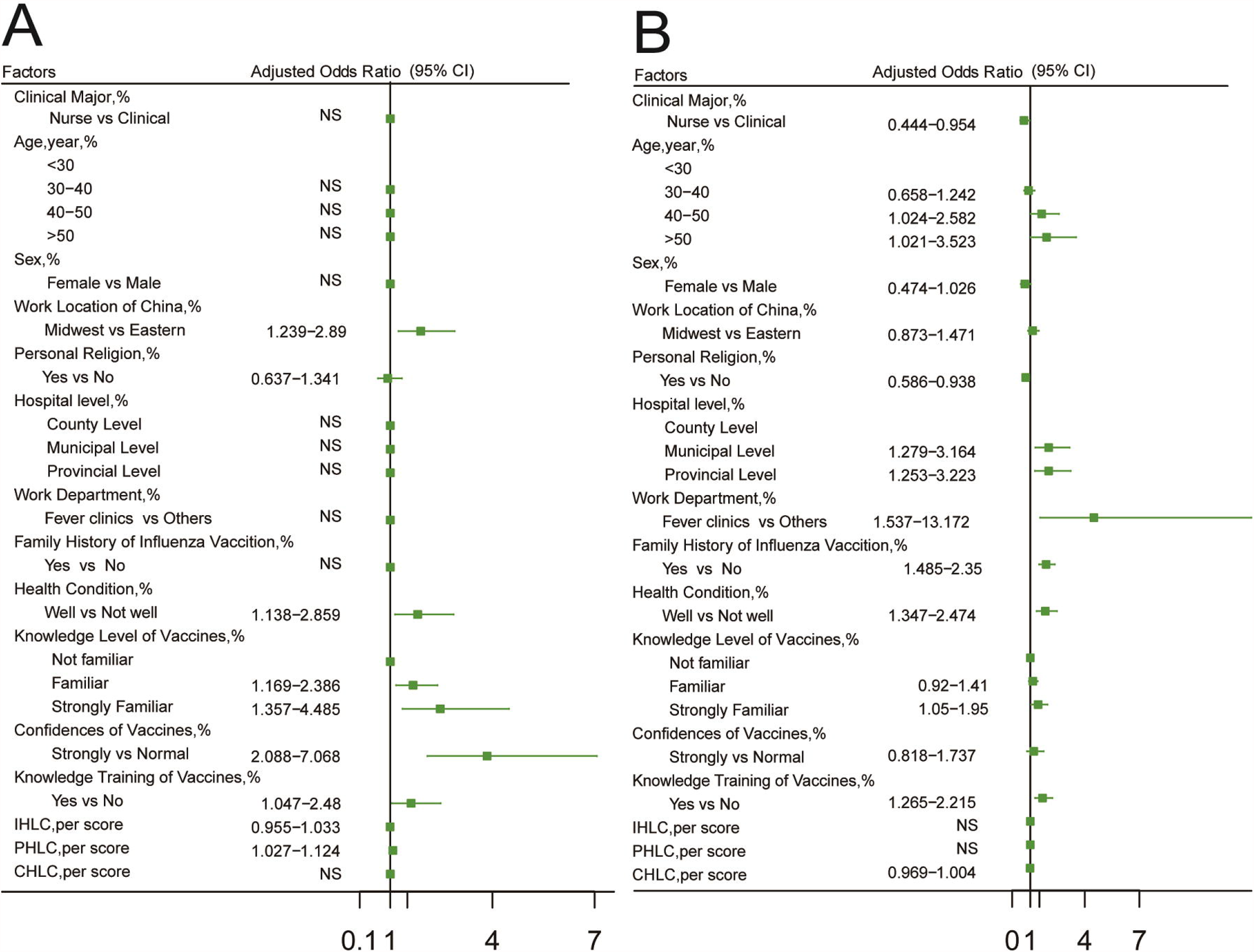

As shown in Figure 1, the vaccination willingness was significantly higher if the HCWs were working in midwestern China, had been trained with the knowledge of vaccines, had strong familiarity with vaccines, had more confidence in vaccines, and had healthy physical condition. Comparatively, subjects with older age (30–40 years vs. less than 30 years, OR=1.626, 95%CI=1.024-2.582) and 40–50 years vs. 30 years, OR=1.896, 95%CI=1.021-3.523), had healthy condition, working in the fever department, and working in higher hospital level (municipal vs. county OR=2.012, 95%CI=1.279-3.164 provincial vs. county, OR=2.01, 95%CI=1.253-3.223) presented higher vaccination rates in the survey. Subjects with religious beliefs, working as a nurse, had been trained with the knowledge of vaccines and were more familiar with the vaccine had lower vaccination rates. A history of influenza vaccination was also positively associated with higher vaccination rates among the HCWs (Table A2 and Figure. 1).

### Subjective opinions on COVID-19 vaccination among HCWs

To better understand the actual concerns of HCWs and to improve their willingness to vaccinate, subjective reasons related to COVID-19 vaccination were explored in the study population. Figure 2 shows the main reasons why HCWs would accept the vaccine, and, the top five reasons were: they are part of a high-risk group that needs to be vaccinated; they feel responsibility for reducing COVID-19 cases; they want to support national vaccine management; it is a recommendation by the government; safety and effectiveness of the vaccine.

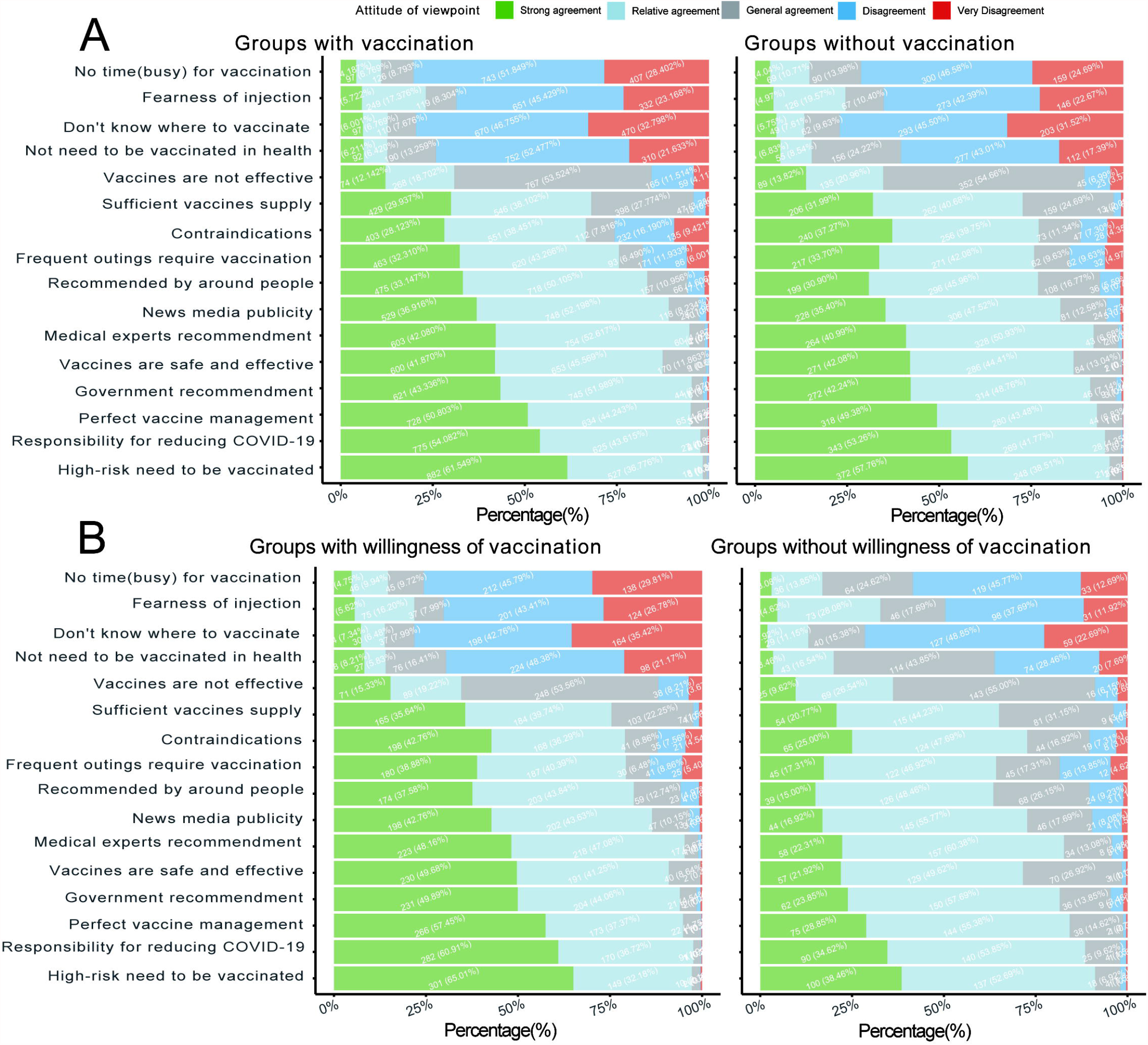

### Adverse effects

Of the 1433 people that have been vaccinated, 673 (47.0%) had one dose and 760 (53.0%) had two doses; 1422 (99.2%) chose a domestic vaccine and 11 (0.8%) chose an imported vaccine. A total of 135 adverse effects (9.4%) were reported, including weakness (74, 5.2%) and headache/dizziness (58, 4.0%) (Figure. 3).

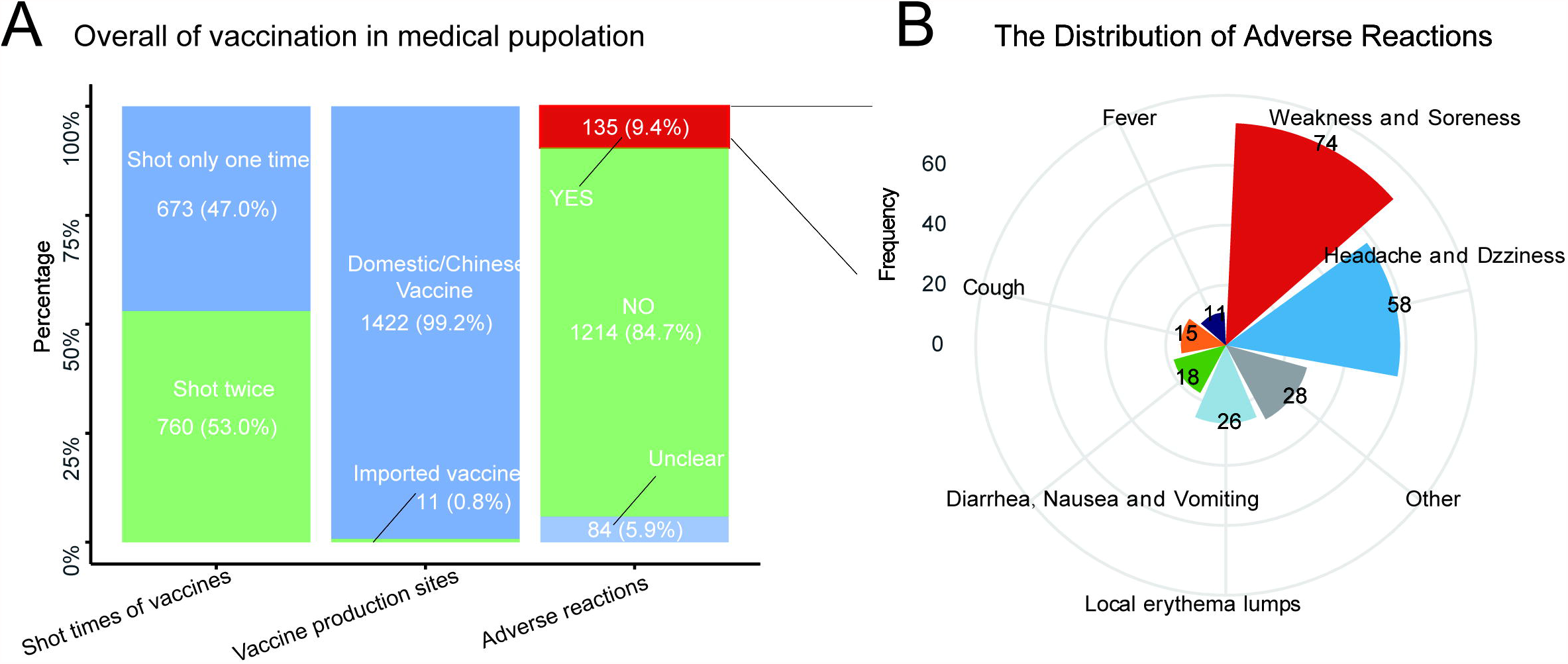

## Discussion

This study firstly provides an in-depth analysis of determinants for vaccination acceptance among HCWs from 21 provinces in China. Of the 2156 participants we included; the vaccination rate was 66.5%. A higher vaccination rate was associated with personal characteristics (male participant, older age, work as clinician, no personal religion, bachelor degree and higher, and healthy physical condition), working environment (longer years of clinical work, working in midwestern China, working in a fever clinic), and more familiarity and belief in the vaccine. Of the 723 unvaccinated participants, 10.9% were unwilling or very unwilling to receive the vaccination. Strong willingness to take the vaccine was related to having a healthy physical condition, considerable knowledge of the vaccine, and strong confidence in the vaccine, which are consistent with published literature.^22,28-30^ Moreover, personal religion and obtained training on the vaccine were also associated with self-reported willingness to receive it.

The MHLC results suggest that willingness to receive the vaccine is primarily influenced by powerful others’ actions. Multivariate analyses show that people’s willingness to receive the vaccine was significantly related to their confidence, familiarity, and training on the vaccine, which may be a consequence of the national CDC’s endorsement. A survey in China showed that the public’s willingness of vaccination could increase from 62.53% to 85.82% if clinicians recommended it.^31^ Similarly, an American survey reported a higher probability of accepting the vaccine if it was endorsed by the CDC of America (coefficient 0.09, 95%CI: 0.07-0.11) and by the WHO (coefficient 0.06, 95%CI: 0.04-0.08). These findings highlight the importance of national CDC and healthcare agencies when promoting vaccination and other health activities.

Moreover, the results show that the willingness to vaccinate was stronger in HCWs from midwestern regions than those from eastern regions. Considering that the vaccines are equally and sufficiently distributed in each province across the country,^32^ this difference might reflect the comparatively weaker healthcare system in the midwestern regions of China,^33^ specifically, people working in the midwest may be more worried about the result if they are infected, and, consequently, are more willing to be vaccinated. While acceptance of the vaccine was associated with the working location, the imbalanced acceptance rate across the country might be due to the imbalance of medical resources in different regions of China,^34^ reflecting that efficient delivery of high-quality healthcare to each province is vital for China’s future medical development.

Interestingly, it was found that self-reported willingness to receive the vaccine may not correlated with taking the vaccine. Whilst vaccination willingness did not differ among different age groups, the actual vaccination rate was significantly higher in people aged ≥40 years, which is consistent with the findings in other countries.^22,35-38^ This presumably because the immune function decreases with age and the incidence and mortality of COVID-19 are relatively higher in older adults.^39^ Furthermore, young people often do not have a strong demand for vaccines and tend to adopt a wait-and-see attitude. This attitude was likely heightened because the pandemic was effectively controlled in China. Hence, the vaccination behavior of younger individuals was observed to be less than the elderly. A survey conducted among young people and medical students also found a lack of preventive attitudes when facing the COVID-19 epidemic.^40,41^ Lazarus et al. reported that people ≥50 were significantly more favorably disposed to vaccination than younger participants in Canada, Poland, France, Germany, Sweden, and the UK, but not in China,^42^ which contrasts with the results of the present study. This may be because at the time of Lazarus’ analysis, elderly people were not recommended to be vaccinated in China, since the safety of the Chinese vaccine for people >60 years old had not been confirmed at that time. However, the CDC of China currently recommends vaccination for elderly people in light of increasing evidence about its safety and effectiveness.^43,44^ However, further investigation is needed to confirm this finding in larger clinical trials.

We observed that HCWs from fever clinics were more likely to be vaccinated, whilst most HCWs believed that high-risk groups should have priority. Moreover, Nguyen et al. found that the acceptance of the seasonal influenza vaccine was related to the fear of getting infected (66%).^45^ Given these, the mortality and infectivity of the virus might be influencing factors for vaccine acceptance. The participants in this study are HCWs who have better knowledge of the SARS-CoV-2 virus compared to the general public, and, consequently, the vaccination rate and intentions were higher. This shows the importance of raising public’s awareness of the SARS-CoV-2 virus when promoting the vaccination throughout the country. The authorities may also need to start educational campaigns much earlier in future public health emergencies.

As reported by the previous research, the most common reason for vaccination resistance was concern about its side effects.^22,28,46^ One study showed that the adverse reactions of the SARS-CoV-2 vaccine are similar to those of the influenza vaccine after vaccination^47^ and the normal and systemic reaction rates for the influenza vaccine are 2.7% and 3.0%, respectively.^48^ This may help to explain why subjects in this study who reported self or family history of influenza vaccination were more likely to be vaccinated, since they are more familiar with the potential side effects of the SARS-CoV-2 vaccine. Furthermore, Nguyen et al. showed that non-physicians may be more concerned about the vaccine’s safety than physicians,^45^ suggesting that the general public may be more worried about the vaccine due to their lack of knowledge. Therefore, healthcare agencies need to increase vaccine-related education to the general public, particularly on: (1) the development and manufacturing processes for vaccines; (2) the similarities between the seasonal influenza vaccine and the SARS-CoV-2 vaccine; and (3) the efficiency and safety of vaccines based on the latest clinical trials. Moreover, authorities should strive to publish the true reason for side effects, which could help to distinguish legitimate safety concerns from events that are temporally associated with but not caused by vaccination. The inappropriate assessment of vaccine safety data can severely undermine the acceptance of the vaccine, and, consequently, influence the success of a mass vaccine campaign.^49,50^

This study has certain limitations. Firstly, at the completion of this survey, China has not recommended the vaccination for people over 60 years old. The vaccination status and associated factors among HCWs in this age group could not be analyzed. However, as most HCWs in China retire when they reach 60, the population in this study is likely to represent HCWs who were working at hospitals during the time of data collection. Secondly, we only conducted a cross-sectional multivariate analysis of the survey data, which can only show the correlation between each factor and vaccination willingness and vaccination behavior, but it cannot prove its causality; therefore, further longitudinal studies are necessary. Finally, as this study was based on self-reported data, it has certain weaknesses that may serve as sources of bias in data interpretation. Despite these limitations, the large sample size of this study and the representative demographics of Chinese HCWs provides relevant information on the vaccination status of HCWs and a reference for the subsequent formulation of vaccination policies.

## Conclusions

Protecting HCWs against COVID-19 is crucial for maintaining the efficacy of the healthcare system during the pandemic. This study suggests that the characteristics of HCWs, working environment, and familiarity and confidence of the vaccine were related to the self-reported willingness to receive the vaccine. Results of this study can not only help to formulate pertinent policies and increase vaccination coverage; they may also provide instructions for future public health emergencies.

## Supporting information

conflict of interest

## Data Availability

Data in the manuscript will not be open, unless it is allowed under the corresponding author's permit

## Acknowledgments

Conceptualization, X.Y. and P.S.; methodology, X.Y., W.Y., J.Y. and Z.R.; software, X.Y. and Y.G.; formal analysis, X.Y. and Y.G.; investigation, L.C., A.D., Q.Y., C.Z., Y.L., Y.W., and S.H.; writing—original draft preparation, X.Y., W.Y. and J.Y.; writing—review and editing, X.Y., W.Y., J.Y., Y.G., Z.R., L.C., A.D., Q.Y., C.Z., Y.L., Y.W., S.H. and P.S. All authors have read and agreed to the published version of the manuscript.

## Institutional Review Board Statement

The study was conducted according to the guidelines of the Declaration of Helsinki, and approved by the Ethics Committee of Xiamen Medical College and passed the audit of China Clinical Trial Registration Center (Registration number: ChiCTR2100042804).

## Informed Consent Statement

Informed consent was obtained from all subjects involved in the study.

