## Supplementary material for "The landscape of COVID-19 vaccination among healthcare workers at the first round of COVID-19 vaccination in China: willingness, acceptance and self-reported adverse effects": conflict of interest

**Figure A1**: The flowchart of the present study

**Figure A2**: The sources for obtaining vaccine-related information

**Figure A3**: MHLC psychology results for the participants

**Table A1**: The characteristics of including Subjects for Vaccines in the Survey

**Table A2**: Univariate Logistic Regression Analysis for Willingness and Whether Receive to Vaccines in the including subjects

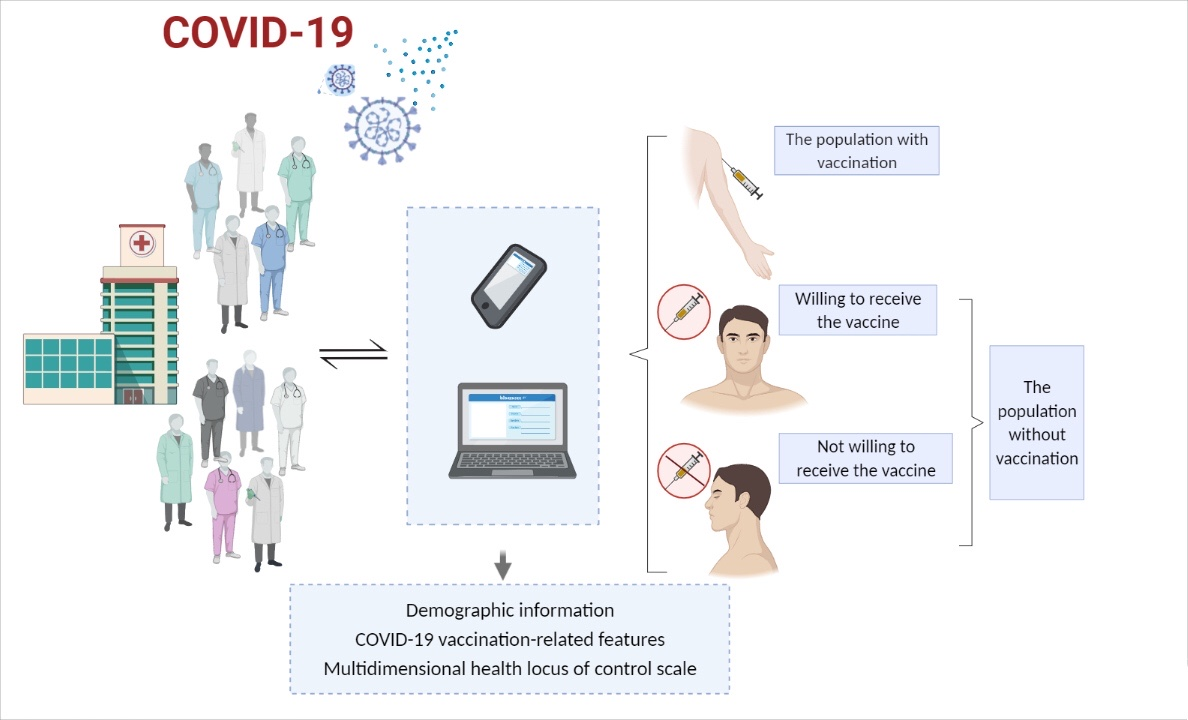

**Figure A1.** The flowchart of the present study. The figure was generated using the Biorender software.

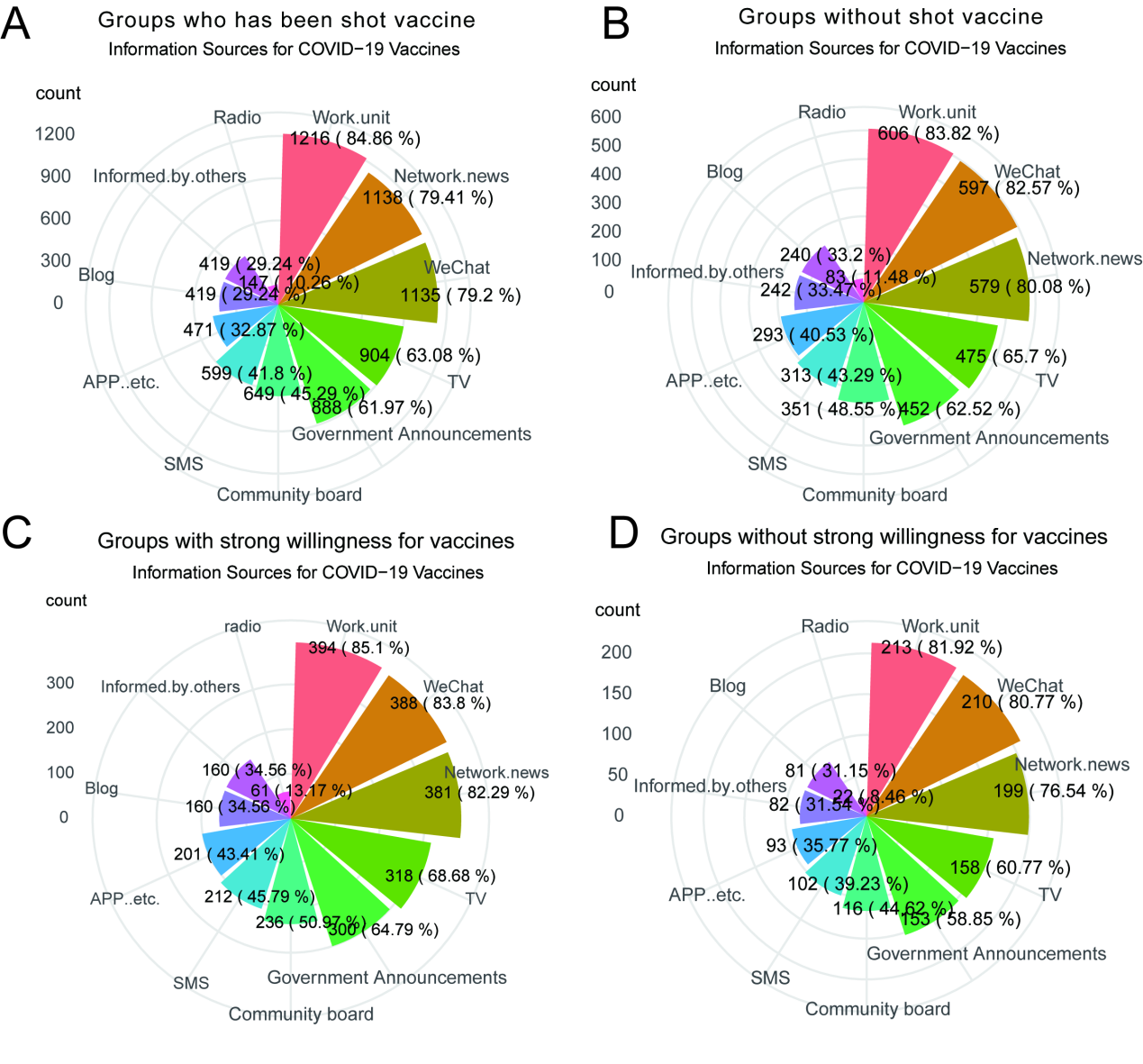

**Figure A2.** The sources for obtaining vaccine-related information.

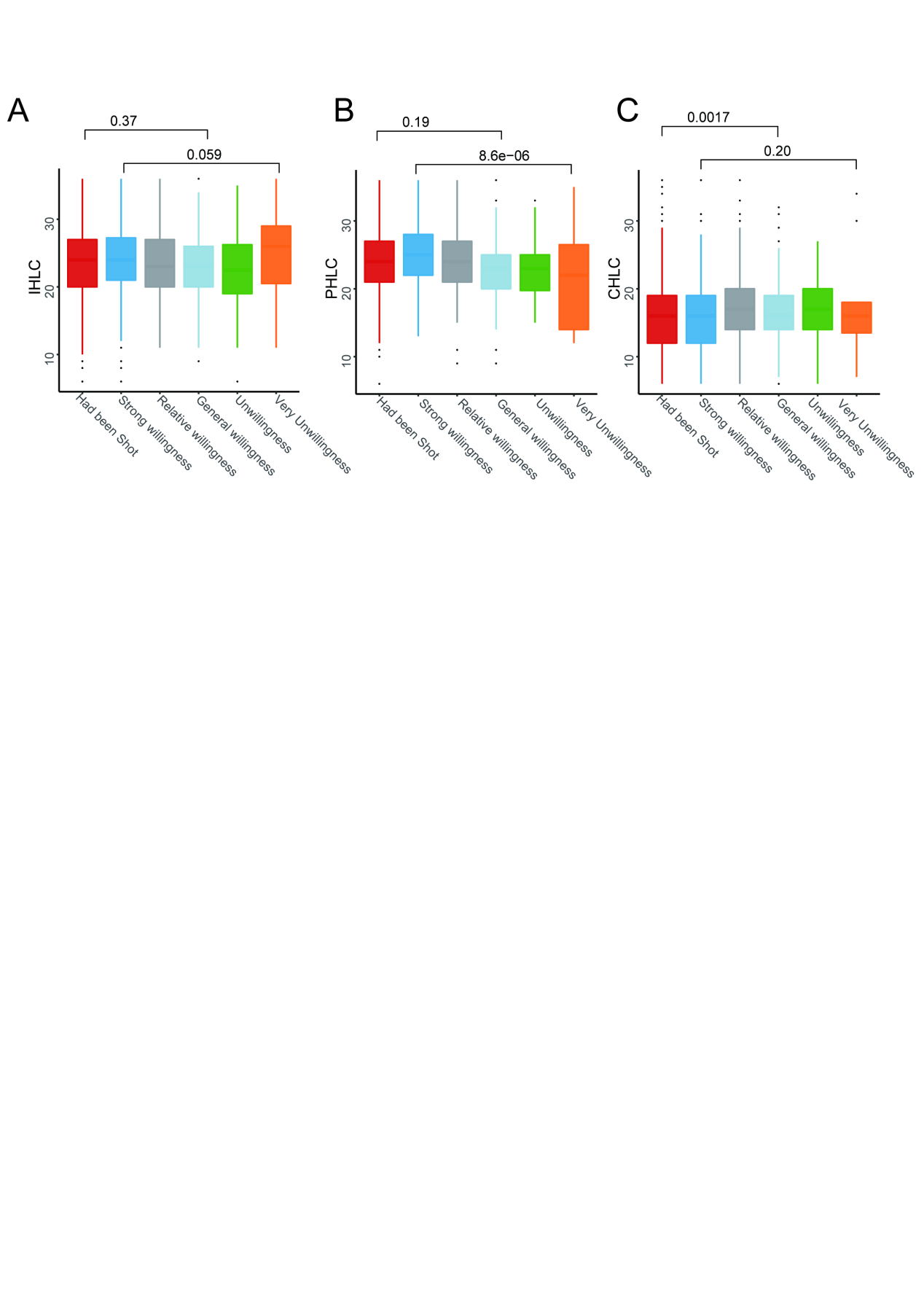
**Figure A3.** MHLC psychology results for the participants.**A,** IHLC:internal health locus of control. **B.**PHLC: powerful other’s health locus of control. **C.**CHLC: chance health locus of control.

**Table A1.** The comparisons in different groups of Willingness and Receiving of vaccines in population.

| Characteristic and category | | Receive Vaccine(n=1433) | Not Receive Vaccine(n=723) | *P1* | Willingness to receive vaccine | | | | | *P2* |
| --- | --- | --- | --- | --- | --- | --- | --- | --- | --- | --- |
|  |  |  |  |  | Strong Willingness(n=228) | Relative Willingness(n=235) | General Willingness(n=181) | Unwillingness(n=68) | Very Unwillingness(n=11) |  |
| Demographic Characteristics | | | | | | | | | | |
| Sex, n(%) | | | | | | | | | | |
|  | Men | 203(14.2) | 57(7.9) | **<0.001** | 24(10.5) | 14(6.0) | 13(7.2) | 4(5.9) | 2(18.2) | 0.24 |
|  | Women | 1230(85.8) | 666(92.1) |  | 204(89.5) | 221(94.0) | 168(92.8) | 64(94.1) | 9(81.8) |  |
| Age (year, mean (SD^a^)) | | 33.86(8.65) | 31.04(7.19) | **<0.001** | 31.06(6.68) | 31.29(7.20) | 30.87(7.63) | 30.62(7.57) | 30.64(8.36) | 0.96 |
| Clinical occupation, n(%) | | | | | | | | | | |
|  | Clinical doctor | 271(18.9) | 72(10.0) | **<0.001** | 21(9.2) | 27(11.5) | 16(8.8) | 5(7.4) | 3(27.3) | 0.27 |
|  | Nurse | 1162(81.1) | 651(90.0) |  | 207(90.8) | 208(88.5) | 165(91.2) | 63(92.6) | 8(72.7) |  |
| Race, n(%) | | | | | | | | | | |
|  | Han | 1391(97.1) | 696(96.3) | 0.38 | 217(95.2) | 226(96.2) | 176(97.2) | 66(97.1) | 11(100.0) | 0.78 |
|  | Others | 42(2.9) | 27(3.7) |  | 11(4.8) | 9(3.8) | 5(2.8) | 2(2.9) | 0(0.0) |  |
| Personal religion, n(%) | | | | | | | | | | |
|  | No | 1062(74.1) | 458(63.3) | **<0.001** | 158(69.3) | 148(63.0) | 108(59.7) | 34(50.0) | 10(90.9) | **0.01** |
|  | Yes | 371(25.9) | 265(36.7) |  | 70(30.7) | 87(37.0) | 73(40.3) | 34(50.0) | 1(9.1) |  |
| Education, n(%) | | | | | | | | | | |
|  | College or under | 487(34.0) | 324(44.8) | **<0.001** | 88(38.6) | 106(45.1) | 88(48.6) | 36(52.9) | 6(54.5) | 0.27 |
|  | Bachelor | 868(60.6) | 373(51.6) |  | 134(58.8) | 120(51.1) | 86(47.5) | 29(42.6) | 4(36.4) |  |
|  | Master or above | 78(5.4) | 26(3.6) |  | 6(2.6) | 9(3.8) | 7(3.9) | 3(4.4) | 1(9.1) |  |
| Average monthly income of family, CNY^b^, n(%) | | | | | | | | | | |
|  | ≤2000 | 61(4.3) | 43(5.9) | 0.23 | 14(6.1) | 11(4.7) | 10(5.5) | 6(8.8) | 2(18.2) | 0.20 |
|  | 2000-4000 | 540(37.7) | 279(38.6) |  | 90(39.5) | 98(41.7) | 61(33.7) | 25(36.8) | 5(45.5) |  |
|  | 4000-6000 | 439(30.6) | 225(31.1) |  | 72(31.6) | 79(33.6) | 56(30.9) | 16(23.5) | 2(18.2) |  |
|  | 6000-8000 | 200(14.0) | 82(11.3) |  | 25(11.0) | 24(10.2) | 27(14.9) | 5(7.4) | 1(9.1) |  |
|  | ≥8000 | 193(13.5) | 94(13.0) |  | 27(11.8) | 23(9.8) | 27(14.9) | 16(23.5) | 1(9.1) |  |
| Health condition, n(%) | | | | | | | | | | |
|  | Not well | 118(8.2) | 103(14.2) | <0.001 | 24(10.5) | 25(10.6) | 37(20.4) | 15(22.1) | 2(18.2) | **0.006** |
|  | Well | 1315(91.8) | 620(85.8) |  | 204(89.5) | 210(89.4) | 144(79.6) | 53(77.9) | 9(81.8) |  |
| Work Environment | | | | | | | | | | |
| Time of employment, n(%) | | | | | | | | | | |
|  | ≤1years | 75(5.2) | 49(6.8) | **<0.001** | 15(6.6) | 17(7.2) | 10(5.5) | 6(8.8) | 1(9.1) | 0.24 |
|  | 1-2years | 124(8.7) | 50(6.9) |  | 10(4.4) | 14(6.0) | 21(11.6) | 4(5.9) | 1(9.1) |  |
|  | 3-5years | 211(14.7) | 150(20.7) |  | 51(22.4) | 40(17.0) | 38(21.0) | 19(27.9) | 2(18.2) |  |
|  | 6-9years | 279(19.5) | 190(26.3) |  | 61(26.8) | 75(31.9) | 40(22.1) | 12(17.6) | 2(18.2) |  |
|  | ≥10years | 744(51.9) | 284(39.3) |  | 91(39.9) | 89(37.9) | 72(39.8) | 27(39.7) | 5(45.5) |  |
| Work location, n(%) | | | | | | | | | | |
|  | Eastern | 799(55.8) | 492(68.0) | **<0.001** | 132(57.9) | 161(68.5) | 142(78.5) | 51(75.0) | 6(54.5) | **<0.001** |
|  | Midwestern | 634(44.2) | 231(32.0) |  | 96(42.1) | 74(31.5) | 39(21.5) | 17(25.0) | 5(45.5) |  |
| Risk level of location, n(%) | | | | | | | | | | |
|  | High | 0(0.0) | 1(0.1) | 0.20 | 1(0.4) | 0(0.0) | 0(0.0) | 0(0.0) | 0(0.0) | 0.44 |
|  | Median | 11(0.8) | 9(1.2) |  | 2(0.9) | 3(1.3) | 2(1.1) | 1(1.5) | 1(9.1) |  |
|  | Low | 1422(99.2) | 713(98.6) |  | 225(98.7) | 232(98.7) | 179(98.9) | 67(98.5) | 10(90.9) |  |
| Clinical department, n(%) | | | | | | | | | | |
|  | Fever clinics | 30(2.1) | 4(0.6) | **0.01** | 0(0.0) | 2(0.9) | 2(1.1) | 0(0.0) | 0(0.0) | 0.54 |
|  | Others | 1403(97.9) | 719(99.4) |  | 228(100.0) | 233(99.1) | 179(98.9) | 68(100.0) | 11(100.0) |  |
| Hospital level, n(%) | | | | | | | | | | |
|  | County level | 47(3.3) | 48(6.6) | **<0.001** | 10(4.4) | 19(8.1) | 13(7.2) | 5(7.4) | 1(9.1) | 0.68 |
|  | Municipal level | 728(50.8) | 389(53.8) |  | 123(53.9) | 119(50.6) | 101(55.8) | 41(60.3) | 5(45.5) |  |
|  | Provincial level | 658(45.9) | 286(39.6) |  | 95(41.7) | 97(41.3) | 67(37.0) | 22(32.4) | 5(45.5) |  |
| Familiarity and Beliefs for Vaccine | | | | | | | | | | |
| Family history for influenza vaccination, n(%) | | | | | | | | | | |
|  | Yes | 485(33.8) | 140(19.4) | **<0.001** | 53(23.2) | 42(17.9) | 29(16.0) | 15(22.1) | 1(9.1) | 0.52 |
|  | No | 855(59.7) | 514(71.1) |  | 153(67.1) | 172(73.2) | 134(74.0) | 45(66.2) | 10(90.9) |  |
|  | Unknown | 93(6.5) | 69(9.5) |  | 22(9.6) | 21(8.9) | 18(9.9) | 8(11.8) | 0(0.0) |  |
| Familiarity of COVID-19 vaccine, n(%) | | | | | | | | | | |
|  | Comprehensive knowledge | 245(17.1) | 86(11.9) | **<0.001** | 47(20.6) | 20(8.5) | 15(8.3) | 3(4.4) | 1(9.1) | **<0.001** |
|  | Sufficient knowledge | 735(51.3) | 352(48.7) |  | 125(54.8) | 120(51.1) | 80(44.2) | 23(33.8) | 4(36.4) |  |
|  | General knowledge | 415(29.0) | 242(33.5) |  | 52(22.8) | 83(35.3) | 74(40.9) | 29(42.6) | 4(36.4) |  |
|  | Limited knowledge | 35(2.4) | 42(5.8) |  | 4(1.8) | 12(5.1) | 12(6.6) | 12(17.6) | 2(18.2) |  |
|  | Not known at all | 3(0.2) | 1(0.1) |  | 0(0.0) | 0(0.0) | 0(0.0) | 1(1.5) | 0(0.0) |  |
| Obtained training on vaccine, n(%) | | | | | | | | | | |
|  | Yes | 1279(89.3) | 594(82.2) | **<0.001** | 201(88.2) | 195(83.0) | 135(74.6) | 55(80.9) | 8(72.7) | **0.03** |
|  | No | 139(9.7) | 117(16.2) |  | 26(11.4) | 34(14.5) | 43(23.8) | 11(16.2) | 3(27.3) |  |
|  | Unknown | 15(1.0) | 12(1.7) |  | 1(0.4) | 6(2.6) | 3(1.7) | 2(2.9) | 0(0.0) |  |
| Confidence of vaccines, n(%) | | | | | | | | | | |
|  | Yes | 813(56.7) | 362(50.1) | **0.003** | 172(75.4) | 117(49.8) | 50(27.6) | 18(26.5) | 5(45.5) | **<0.001** |
|  | No | 542(37.8) | 301(41.6) |  | 46(20.2) | 109(46.4) | 112(61.9) | 32(47.1) | 2(18.2) |  |
|  | Unknown | 78(5.4) | 60(8.3) |  | 10(4.4) | 9(3.8) | 19(10.5) | 18(26.5) | 4(36.4) |  |
| IHLC^c^ (mean(SD)) | | 23.64(5.28) | 23.42(5.29) | 0.35 | 24.20(5.86) | 23.31(4.88) | 22.87(4.60) | 22.46(5.64) | 24.55(8.08) | 0.045 |
| PHLC^d^ (mean(SD)) | | 24.11(4.74) | 23.84(4.69) | 0.20 | 25.01(4.75) | 23.92(4.51) | 22.76(4.48) | 22.85(4.05) | 21.45(7.98) | <0.001 |
| CHLC^e^ (mean(SD)) | | 16.04(5.36) | 16.63(5.26) | 0.02 | 16.30(6.20) | 16.93(5.00) | 16.48(4.19) | 16.97(4.79) | 17.45(8.08) | 0.68 |

^a^SD: standard deviation.

^b^CNY: Chinese Yuan.

^c^IHLC: internal health locus of control.

dPHLC: powerful other’s health locus of control.

eCHLC: chance health locus of control.

**Table A2.**Univariate Logistic Regression Analysis for Willingness and Whether Receive to Vaccines in the including subjects.

| Item | | willingness to receive vaccine | | | receive vaccine vs not receive | | |
| --- | --- | --- | --- | --- | --- | --- | --- |
|  |  | OR^a^ | 95%CI | P-value | OR | 95%CI | P-value |
| Age | | | | | | | |
|  | <30 (ref^b^) | 1.0 |  |  | 1.0 |  |  |
|  | 30-40 | 1.26 | 0.91-1.74 | 0.17 | 1.23 | 1.01-1.49 | 0.04 |
|  | 40-50 | 0.99 | 0.57-1.71 | 0.97 | 2.43 | 1.79-3.30 | <0.001 |
|  | >50 | 1.24 | 0.49-3.15 | 0.65 | 2.91 | 1.78-4.76 | <0.001 |
| Sex | | | | | | | |
|  | Male (ref) | 1.0 |  |  | 1.0 |  |  |
|  | Female | 0.88 | 0.50-1.56 | 0.67 | 0.52 | 0.38-0.71 | <0.001 |
| Clinical occupation | | | | | | | |
|  | Clinical Doctor (ref) | 1.0 |  |  | 1.0 |  |  |
|  | Nurse | 0.88 | 0.53-1.47 | 0.63 | 0.47 | 0.36-0.63 | <0.001 |
| Race | | | | | | | |
|  | Han (ref) | 1.0 |  |  | 1.0 |  |  |
|  | Others | 1.63 | 0.68-3.91 | 0.27 | 0.78 | 0.48-1.27 | 0.33 |
| Personal religion | | | | | | | |
|  | No (ref) | 1.0 |  |  | 1.0 |  |  |
|  | Yes | 0.72 | 0.53-0.99 | 0.04 | 0.60 | 0.50-0.73 | <0.001 |
| Education | | | | | | | |
|  | College or under (ref) | 1.0 |  |  | 1.0 |  |  |
|  | Bachelor | 1.43 | 1.05-1.95 | 0.02 | 1.55 | 1.29-1.87 | <0.001 |
|  | Master or above | 0.91 | 0.41-2.05 | 0.83 | 2 | 1.25-3.18 | 0.004 |
| Average monthly income of family, CNY^c^ | | | | | | | |
|  | ≤2000 (ref) | 1.0 |  |  | 1.0 |  |  |
|  | 2000-4000 | 1.49 | 0.77-2.86 | 0.24 | 1.36 | 0.90-2.07 | 0.14 |
|  | 4000-6000 | 1.47 | 0.76-2.86 | 0.26 | 1.38 | 0.90-2.10 | 0.14 |
|  | 6000-8000 | 1.07 | 0.51-2.26 | 0.86 | 1.72 | 1.08-2.74 | 0.02 |
|  | ≥8000 | 0.82 | 0.40-1.70 | 0.59 | 1.45 | 0.91-2.30 | 0.12 |
| Time of employment | | | | | | | |
|  | ≤1 year (ref) | 1.0 |  |  | 1.0 |  |  |
|  | 1-2 years | 0.49 | 0.22-1.10 | 0.08 | 1.62 | 0.995-2.64 | 0.05 |
|  | 3-5 years | 0.819 | 0.42-1.61 | 0.56 | 0.92 | 0.61-1.39 | 0.69 |
|  | 6-9 years | 1.338 | 0.69-2.61 | 0.39 | 0.96 | 0.64-1.44 | 0.84 |
|  | ≥10 years | 0.919 | 0.49-1.74 | 0.80 | 1.71 | 1.17-2.51 | 0.006 |
| Work location | | | | | | | |
|  | Eastern (ref) | 1.0 |  |  | 1.0 |  |  |
|  | Midwest | 1.89 | 1.34-2.67 | 0 | 1.69 | 1.40-2.04 | <0.001 |
| Risk of location | | | | | | | |
|  | No (ref) | 1.0 |  |  | 1.0 |  |  |
|  | Yes | 1.60 | Inf | Inf | 3.50 | Inf | 0.97 |
| Clinical department | | | | | | | |
|  | Others (ref) | 1.0 |  |  | 1.0 |  |  |
|  | Fever clinics | 0.56 | 0.08-4.00 | 0.56 | 3.84 | 1.35-10.94 | 0.01 |
| Hospital level | | | | | | | |
|  | County level (ref) | 1.0 |  |  | 1.0 |  |  |
|  | Municipal level | 1.08 | 0.58-1.99 | 0.81 | 1.91 | 1.25-2.91 | 0.003 |
|  | Provincial level | 1.34 | 0.71-2.51 | 0.36 | 2.35 | 1.54-3.59 | <0.001 |
| Family history for influenza vaccination | | | | | | | |
|  | No (ref) | 1.0 |  |  | 1.0 |  |  |
|  | Yes | 1.23 | 0.83-1.83 | 0.30 | 2.13 | 1.72-2.64 | <0.001 |
| Familiarity of COVID-19 vaccine^d^ | | | | | | | |
|  | Not familiar (ref) | 1.0 |  |  | 1.0 |  |  |
|  | Familiar | 2.03 | 1.47-2.81 | <0.001 | 1.31 | 1.08-1.60 | 0.006 |
|  | Strongly familiar | 3.13 | 1.79-5.48 | <0.001 | 1.79 | 1.35-2.39 | <0.001 |
| Obtained training on vaccine | | | | | | | |
|  | No (ref) | 1.0 |  |  | 1.0 |  |  |
|  | Yes | 1.85 | 1.26-2.72 | 0.002 | 1.8 | 1.40-2.32 | <0.001 |
| Confidence on vaccines | | | | | | | |
|  | Normal (ref) | 1.0 |  |  | 1.0 |  |  |
|  | Strong | 4.38 | 2.48-7.73 | <0.001 | 1.57 | 1.11-2.23 | 0.01 |
| Health condition | | | | | | | |
|  | Not well (ref) | 1.0 |  |  | 1.0 |  |  |
|  | Well | 2.22 | 1.45-3.38 | <0.001 | 1.85 | 1.40-2.45 | <0.001 |
| IHLC^e^ (continuous) | | 1.03 | 1.004-1.06 | 0.03 | 1.01 | 0.99-1.03 | 0.35 |
| PHLC^f^ (continuous) | | 1.09 | 1.05-1.12 | <0.001 | 1.01 | 0.99-1.03 | 0.20 |
| CHLC^g^ (continuous) | | 0.999 | 0.97-1.02 | 0.94 | 0.98 | 0.96-0.995 | 0.02 |

^a^OR: odds ratio.

^b^ref: reference.

^c^CNY: Chinese Yuan.

^d^Items merged for this variable. “Not familiar” refers to people who have limited or no knowledge on vaccine; “familiar” refers to people who have general knowledge on vaccine; “Strongly familiar” refers to people who have comprehensive or sufficient knowledge on vaccine.

^e^IHLC: internal health locus of control.

^f^PHLC: powerful others’ health locus of control.

^g^CHLC: chance health locus of control.
